# Prevalence and clinical presentations of post-COVID-19 conditions in Nepal

**DOI:** 10.1101/2024.04.17.24305977

**Authors:** Lila Bahadur Basnet, Pomawati Thapa, Anup Bastola, Narendra Khanal, Niranjan Panta, Sabin Thapaliya, Melissa Beth Kleine Bingham, Saugat Shrestha, Shital Adhikari, Sudesha Khadka, Priyanka Shrestha, Sadhana Paudel, Kamaraj Arulmozhi Devapitchai

## Abstract

**Introduction:** The COVID-19 pandemic affected millions worldwide. While the major focus was on diagnosis and treatment of the acute phase of the disease, many individuals experienced long-term health even after recovery. These post-COVID-19 conditions encompass a broad range of physical, psychological, and cognitive symptoms that persist beyond the acute phase of the illness. The aim of this study was to find the prevalence and reported presentations of post-COVID-19 conditions in Nepal.

**Method:** This is a retrospective cross-sectional study using structured questionnaire to collect hospital-based data available in the record of Curative Service Division, Department of Health Services of Nepal from October 2021 to September 2022. A total of 6151 cases were recorded in the study. Descriptive analysis was done for demographic and symptoms variable. Association of variables to post-COVID-19 condition are shown using univariate and multivariate logistic regression.

**Results:** Among 6151 respondents (62.25% males), more than half (59.03%) had at least one symptom after recovery from acute COVID-19. The most common symptoms were anxiety (28.5%), loss of appetite (25.3%), shortness of breath (24.13%), fatigue (23.24%), depressed mood (18.79%), muscle ache (17.59%), chest pain (16.81%), headache (14.78%), and palpitation (13%). Multivariate analysis showed increased odds of post-COVID-19 conditions in smokers (adjusted OR [aOR] 1.53), those with chronic lung disease (aOR 1.71), neurological disorder (aOR 2.43) and those with use of supplementary oxygen during acute illness of COVID-19 (aOR 3.37).

**Conclusion:** The prevalence of post-COVID-19 symptoms among the study population is found to be high. The main symptoms from our study and similar studies are anxiety, fatigue, loss of appetite, shortness of breath, muscle pain, headaches. Patients with smoking, hypoxia during acute illness and presence of chronic lung disease and neurological disorder had higher odds of getting post-COVID-19 conditions. Citing the findings, health system should focus upon the management and recording of such conditions for further evidence.

## Introduction

The COVID-19 pandemic affected millions of individuals worldwide causing significant morbidity and mortality. While the immediate focus had been on diagnosing and treating the acute phase of the disease, there is growing recognition of the long-term health consequences experienced by individuals recovering from COVID-19 [1,2]. These post-COVID-19 conditions, commonly referred to as “long COVID” or “post-acute sequelae of SARS-CoV-2 infection” (PASC), encompass a broad range of physical, psychological, and cognitive symptoms that persist beyond the acute phase of the illness [3].

Several studies carried out to understand the features of post-COVID conditions have shown wide range of symptoms affecting the individual’s ability to perform daily activities [4,5]. Though most people who developed COVID-19 fully recovered, approximately 10-20% experience a variety of mid- and long-term effects. The effects of the pandemic, lockdowns, quarantine, and isolation may also have long-term impact. There are no tests or pathognomonic findings to establish the diagnosis of post-COVID-19 conditions. Additionally, there is variability of definition and the terminology used around long-COVID. The definition developed by Delphi consensus by the WHO should help bring uniformity in the research and hence the data related to post-COVID-19 condition [4]. Patients with post-COVID-19 condition require further healthcare services including support group for post-COVID-19 condition has been widely acknowledged for their support and recovery [1]. The identification of the post-COVID-19 conditions and its burden would be the basis for allocation of resources and plan for needs [5].

Few studies have been carried out in a single setting on post-COVID-19 conditions with limited information and evidence on post-COVID-19 conditions across Nepal. This study aimed to find out the prevalence and types of post-COVID symptoms in Nepalese population.

## Method

### Study Design and Setting

A cross-sectional study using secondary data from the electronic record and paper-based record of Curative Service Division (CSD) of Department of Health Services was conducted. CSD hosted the program on post-COVID and lead on the preparation of the treatment protocol for the Management of Post-COVID patients. CSD is also responsible for the provision of basic health services and coordination with hospitals during and beyond the COVID-19 situation.

### Study population

Data (both electronic based and paper based) was recorded between October 2021 through September 22 and included those who recovered from acute illness of COVID-19. The data was accessed on 20^th^ July 2023 from CSD. De-identification of accessed data was done and data with incomplete information were excluded from the study.

### Study tools and techniques

A structured questionnaire was used to collect hospital-based data available in the record of CSD. The structured questionnaire was developed with reference from module 1 and module 2 of WHO Case Report Form (CRF) for post-COVID condition.

### Study Variable

Demographic details, prior comorbidities, symptoms during acute COVID-19, hospitalization and oxygen use during acute COVID-19, symptoms of post-COVID-19 conditions, and history of vaccination against COVID-19 were collected. Fever and cough were excluded in the analysis because it was not clear in the record whether these symptoms reported were present in acute illness or continued after recovery. Post-COVID condition in this study is described the presence of at least one persistent or newly developed symptom one month after recovery from acute illness of COVID-19, in absence of or are not explained by other disease.

### Statistical analysis

The data collected from the records were entered in Excel. Data were analyzed using Stata (version 17). Descriptive analysis was conducted and presented as frequencies and proportions for categorical variables. Associations of post-COVID-19 symptoms with prior comorbidities and various demographic data were analyzed using univariate and multivariate logistic regressions. A p-value of <0.05 was considered statistically significant.

### Ethical consideration

Ethical approval was obtained from National Health Research Council Ethical Review Board. Permission to access data was obtained from the Curative Service Division. The secondary data was used without a personal identifier.

## Results

Of the total 6180 observations screened, 6151 observations were included in the study and the largest number was from the 30-39 years age group. More than half (62.25%) of the individuals were male (Table 1). The prevalence of post-COVID-19 condition (defined here as the presence of at least one symptom one month after recovery from acute COVID) was 59.03%.

**Table 1.** Socio-demographic characteristics of the screened post-COVID-19 cases.

| Variables | Frequency | Percentage |
| --- | --- | --- |
| **Age group** |  |  |
| <=9 years | 31 | 0.5% |
| 10-19 | 140 | 2.3% |
| 20-29 | 1417 | 23.0% |
| 30-39 | 1662 | 27.0% |
| 40-49 | 1094 | 17.8% |
| 50-59 | 797 | 13.0% |
| 60-69 | 566 | 9.2% |
| 70-79 | 309 | 5.0% |
| 80-89 | 126 | 2.0% |
| 90 and above | 9 | 0.1% |
| **Sex** |  |  |
| Male | 3829 | 62.3% |
| Female | 2316 | 37.7% |
| Others | 6 | 0.1% |

As shown in Table 2, individuals had a variety of symptoms, and many had more than one symptom. The most common symptoms were anxiety (28.5%), loss of appetite (25.3%), shortness of breath (24.13%), fatigue (23.24%), depressed mood (18.79%), muscle ache (17.59%), chest pain (16.81%), headache (14.78%), and palpitation (13%). It was found that more than half (59.03%) had at least one symptom after recovery from acute COVID-19.

**Table 2:**
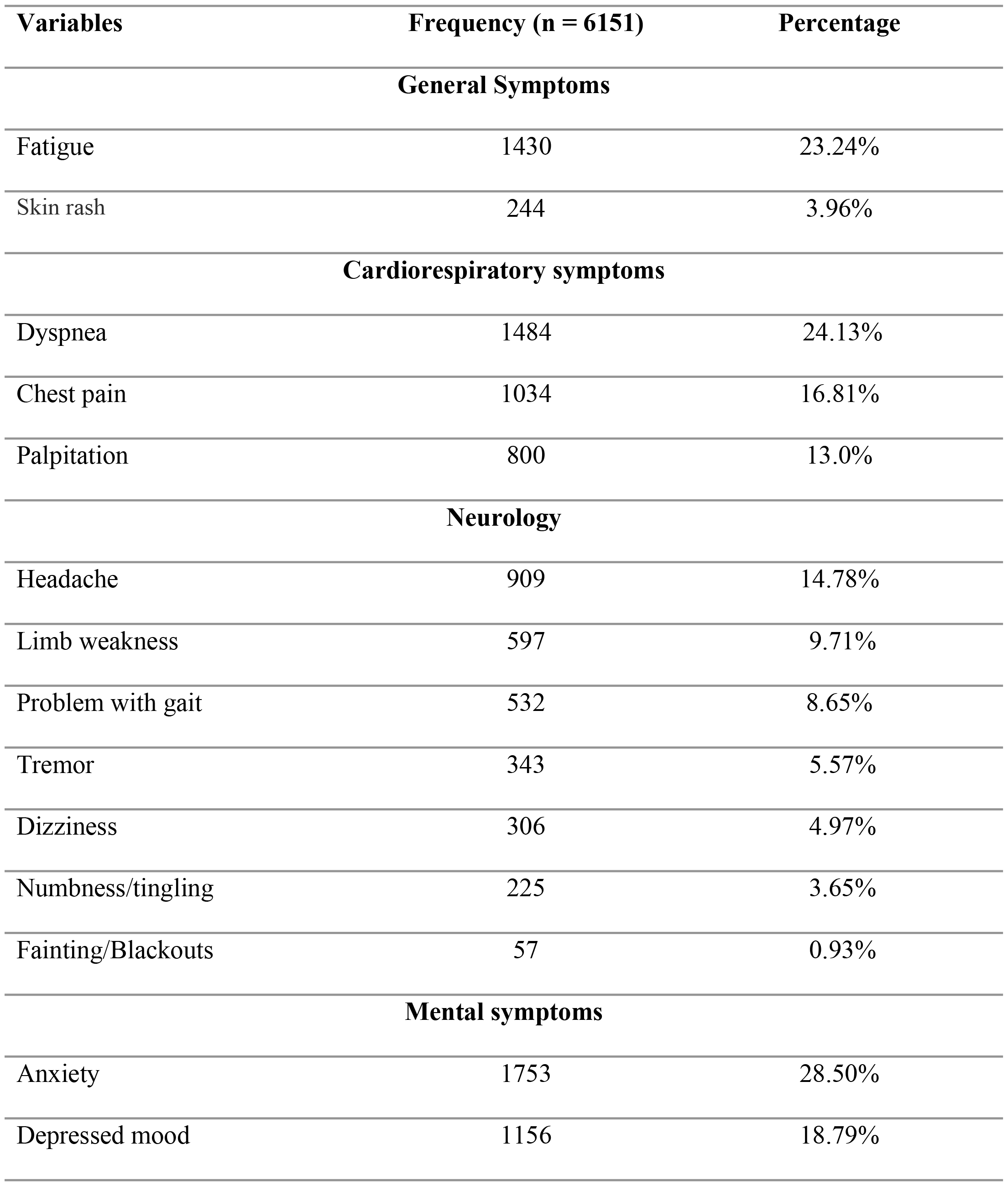
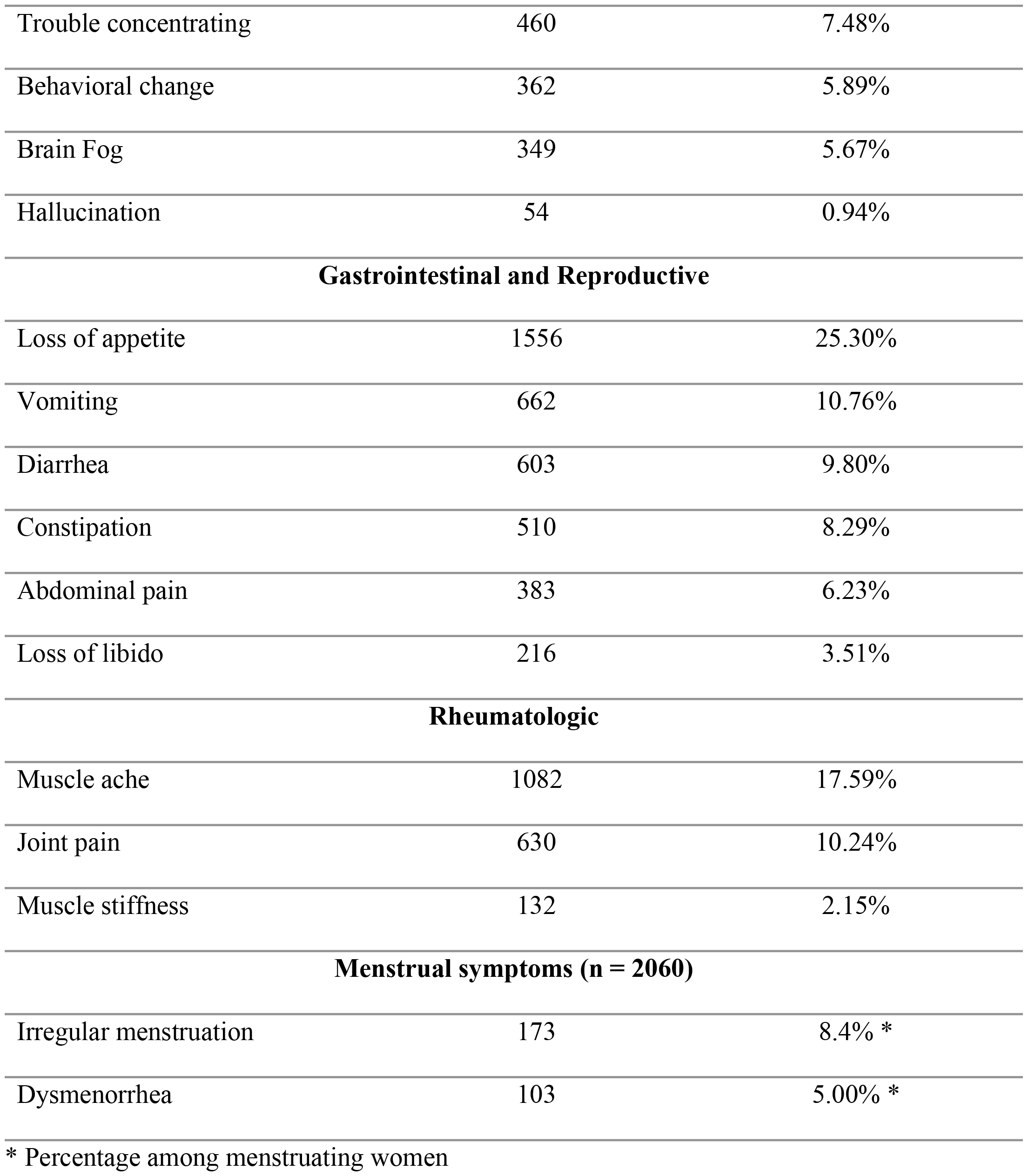
Characteristics of post-COVID-19 conditions (n = 6,151)

| Variables | Frequency (n = 6151) | Percentage |
| --- | --- | --- |
| **General Symptoms** |  |  |
| Fatigue | 1430 | 23.24% |
| Skin rash | 244 | 3.96% |
| **Cardiorespiratory symptoms** |  |  |
| Dyspnea | 1484 | 24.13% |
| Chest pain | 1034 | 16.81% |
| Palpitation | 800 | 13.0% |
| **Neurology** |  |  |
| Headache | 909 | 14.78% |
| Limb weakness | 597 | 9.71% |
| Problem with gait | 532 | 8.65% |
| Tremor | 343 | 5.57% |
| Dizziness | 306 | 4.97% |
| Numbness/tingling | 225 | 3.65% |
| Fainting/Blackouts | 57 | 0.93% |
| **Mental symptoms** |  |  |
| Anxiety | 1753 | 28.50% |
| Depressed mood | 1156 | 18.79% |
| Trouble concentrating | 460 | 7.48% |
| Behavioral change | 362 | 5.89% |
| Brain Fog | 349 | 5.67% |
| Hallucination | 54 | 0.94% |
| **Gastrointestinal and Reproductive** |  |  |
| Loss of appetite | 1556 | 25.30% |
| Vomiting | 662 | 10.76% |
| Diarrhea | 603 | 9.80% |
| Constipation | 510 | 8.29% |
| Abdominal pain | 383 | 6.23% |
| Loss of libido | 216 | 3.51% |
| **Rheumatologic** |  |  |
| Muscle ache | 1082 | 17.59% |
| Joint pain | 630 | 10.24% |
| Muscle stiffness | 132 | 2.15% |
| **Menstrual symptoms (n = 2060)** |  |  |
| Irregular menstruation | 173 | 8.4% * |
| Dysmenorrhea | 103 | 5.00% * |

\* Percentage among menstruating women

Overall, neuropsychiatric symptoms were the most prevalent followed by cardiorespiratory symptoms. The common neuropsychiatric manifestations are shown in Fig 1.

**Fig 1:**
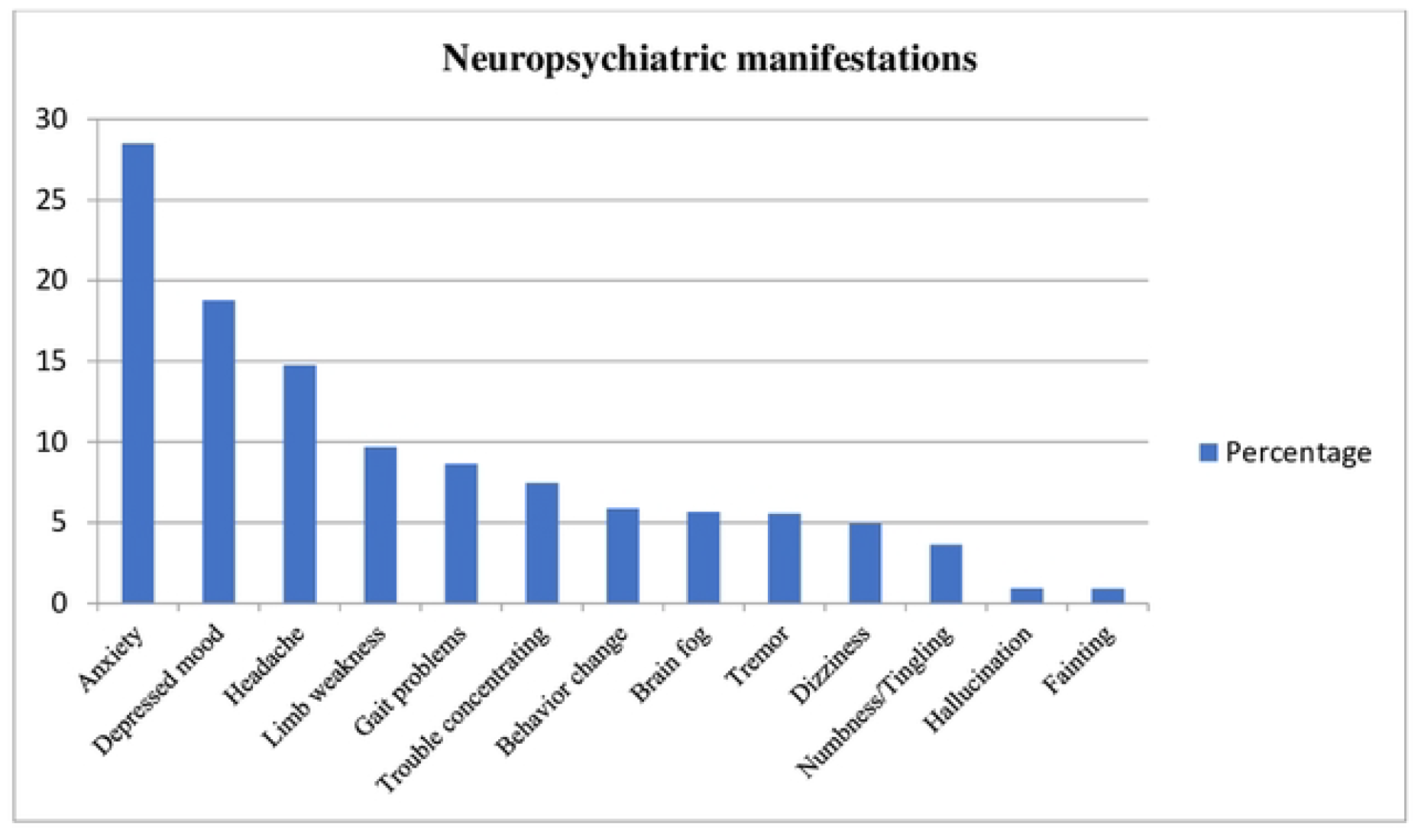
The neuropsychiatric symptoms (shown in percentage)

Smokers (adjusted OR [aOR] 1.53, 95% Confidence Interval [CI] 1.34-1.75), those with chronic lung disease (aOR 1.71, 95% CI 1.22 – 2.39), neurological disorder (aOR 2.43, 95% CI 1.39-4.26), use of supplementary oxygen during acute illness of COVID-19 (aOR 3.37, 95% CI 2.90-3.91) and vaccination (any) against COVID-19 (aOR 1.72, 95% CI 1.53-1.94) had higher odds to get post-COVID-19 condition after recovery from acute illness of COVID-19 even after adjusting for other covariates.(Table 3)

**Table 3:**
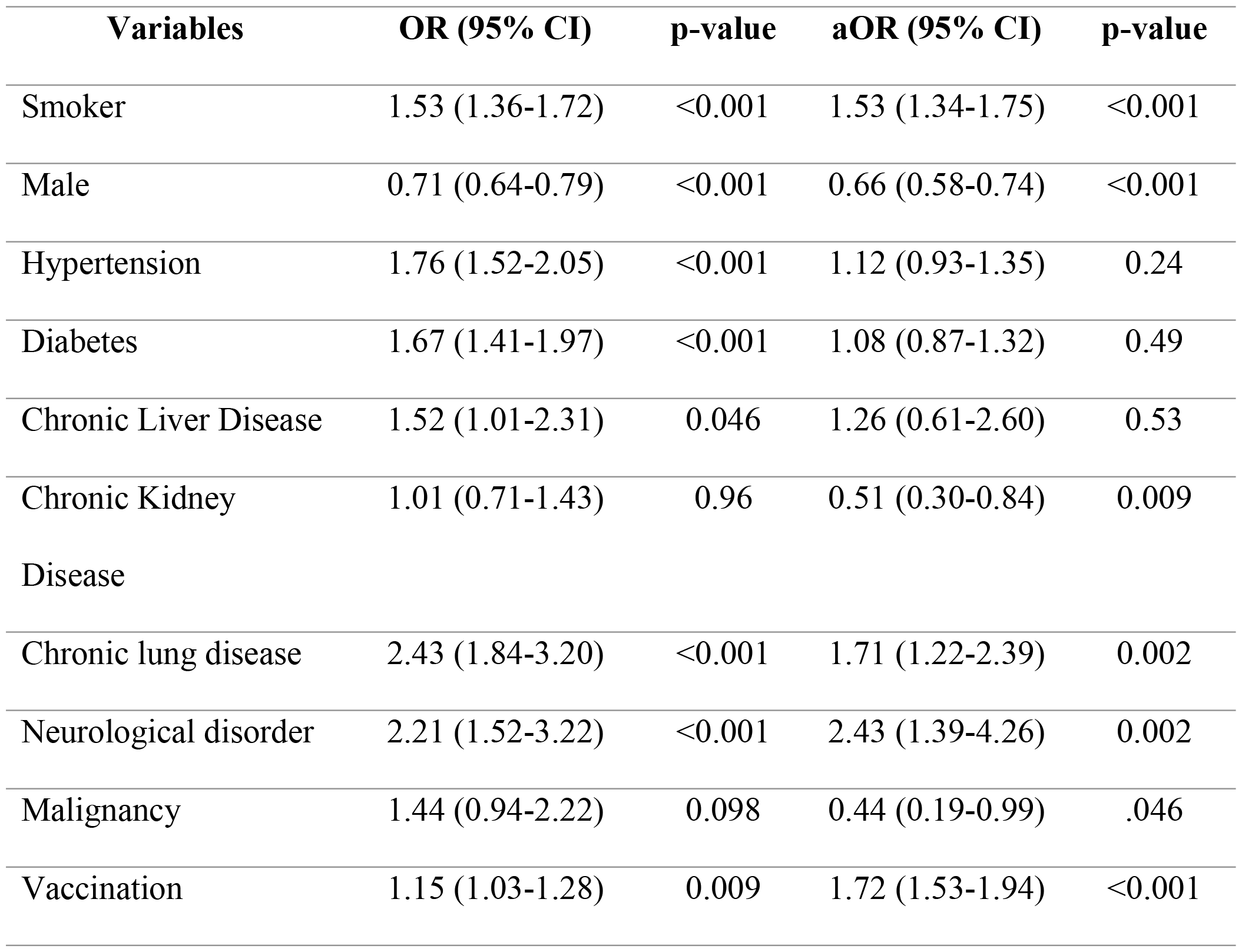
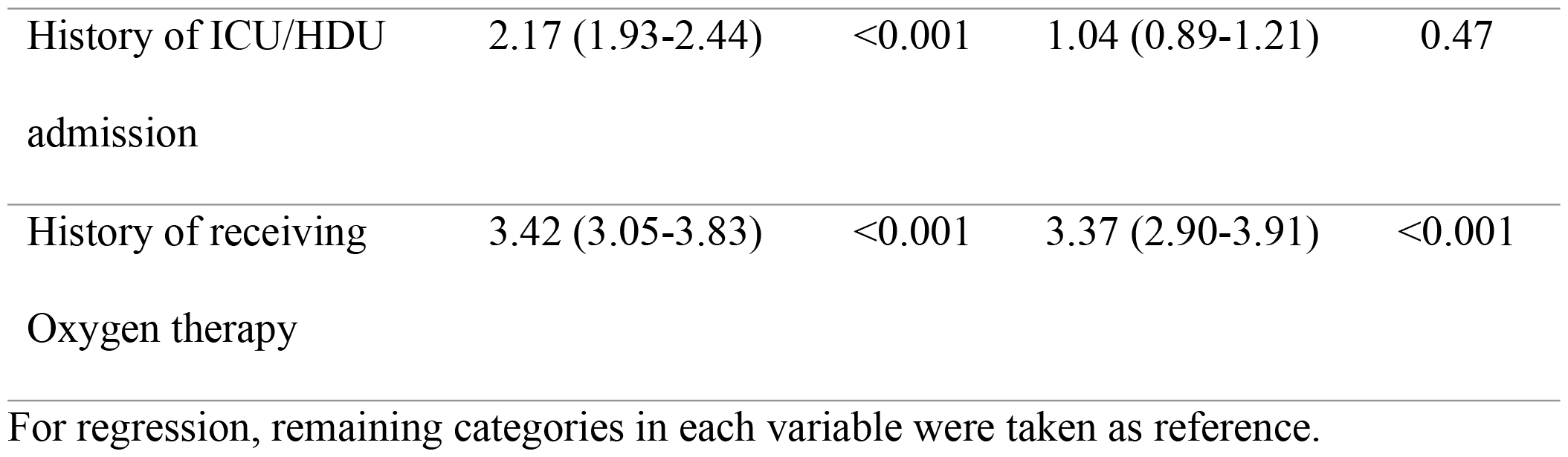
Univariate and multivariate logistic regression for outcome variables.

| Variables | OR (95% CI) | p-value | aOR (95% CI) | p-value |
| --- | --- | --- | --- | --- |
| Smoker | 1.53 (1.36-1.72) | <0.001 | 1.53 (1.34-1.75) | <0.001 |
| Male | 0.71 (0.64-0.79) | <0.001 | 0.66 (0.58-0.74) | <0.001 |
| Hypertension | 1.76 (1.52-2.05) | <0.001 | 1.12 (0.93-1.35) | 0.24 |
| Diabetes | 1.67 (1.41-1.97) | <0.001 | 1.08 (0.87-1.32) | 0.49 |
| Chronic Liver Disease | 1.52 (1.01-2.31) | 0.046 | 1.26 (0.61-2.60) | 0.53 |
| Chronic Kidney Disease | 1.01 (0.71-1.43) | 0.96 | 0.51 (0.30-0.84) | 0.009 |
| Chronic lung disease | 2.43 (1.84-3.20) | <0.001 | 1.71 (1.22-2.39) | 0.002 |
| Neurological disorder | 2.21 (1.52-3.22) | <0.001 | 2.43 (1.39-4.26) | 0.002 |
| Malignancy | 1.44 (0.94-2.22) | 0.098 | 0.44 (0.19-0.99) | .046 |
| Vaccination | 1.15 (1.03-1.28) | 0.009 | 1.72 (1.53-1.94) | <0.001 |
| History of ICU/HDU admission | 2.17 (1.93-2.44) | <0.001 | 1.04 (0.89-1.21) | 0.47 |
| History of receiving Oxygen therapy | 3.42 (3.05-3.83) | <0.001 | 3.37 (2.90-3.91) | <0.001 |

For regression, remaining categories in each variable were taken as reference.

## Discussion

Analyzing the record of 6151 individuals with history of COVID-19 obtained, more than half were found to be symptomatic. Individuals reported plethora of symptoms ranging from anxiety loss of appetite shortness of breath, fatigue, persistent pain in muscles, and persistent headache. This is similar to the findings from a study done in China by Huang Lixue et. al, in which the proportion of patients with at least one sequelae symptom was found to be 68% (831/1227) at 6 months and 49% (620/1272) at 12 months. Furthermore, after one-year individuals with anxiety as a health problem constituted 26 % [331/1271] and those having dyspnea were 30% [380/1271] [2]. Another study done by Maxime Taquet et. al. reports 57% of the patients after whole 6 months had at least one symptom [3].

The prevalence of post-COVID-19 symptoms are being measured worldwide and attempts has been made to quantify the presentations. There are variety of presentations that are difficult to collate and link to the previous COVID-19 status and hence the under-reporting is obvious. An analysis of the data from over 250,000 electronic health records also shows more than 33% presenting with the features of post-COVID-19 condition recorded between 3 and 6 months after a diagnosis of COVID-19 [6].

In a study published by B Anup et. al. from Nepal, dyspnea, fatigue, chest heaviness, and cough were the commonest persistent complaints with 82.2% of the patients presenting with at least one symptom [7].

Similarly, another study conducted among 300 patients in a tertiary center of Nepal shows fatigue as the most common persistent symptom after COVID-19 illness when followed up after 3 months. The other symptoms reported in this study are sore throat, rhinitis, diarrhea, anosmia, ageusia and shortness of breath [8].

Different studies have shown a variety of symptoms that is linked to the previous COVID-19 illness. The duration of follow-up and the severity of illness might play a significant role in the outcomes. Globally, while there has been much effort to establish a common set of symptoms/outcomes for long-COVID, the variance and wide range of reported symptoms makes this challenging [6].

This study also found that smokers, patients with chronic lung disease, neurological disorder and those with use of supplementary oxygen during acute illness of COVID-19 had more prevalence of symptoms. In a systemic review and meta-analysis by Tsampasian V et al comprising 41 studies including 860 783 patients, smoking (OR, 1.10; 95% CI, 1.07-1.13, underlying co-morbidities (OR, 2.48; 95% CI, 1.97-3.13), and hospitalization and ICU admission (OR, 2.37; 95% CI, 2.18-2.56) were found to be the risk factors for development of post-COVID-19 conditions [9]. Multi-regression analysis in their study did not show diabetes and ischemic heart disease increasing the risk of post-COVID-19 conditions as observed in this study.

Males had lower odds of post-COVID-19 symptoms compared to females and this could be because of the inclusion of menstrual problems and also the increased tendency to report non-specific symptoms among females [10]. Similar findings have been reported in a multicenter study conducted in Spain [11].

People vaccinated against COVID had higher odds of post-COVID-19 symptoms than those who were not vaccinated. This contrasts with what has been reported elsewhere [9]. This could be because the symptoms following vaccination are similar to the post-COVID-19 conditions and could have been reported as post-COVID-19 symptoms [9,12]. However, this requires further investigation and was beyond the scope of this study.

Our study points to the need of tracking the patients who were hospitalized for the COVID-19 patients in hospitals of Nepal. As the COVID-19 pandemic placed the hospitals in unprecedented public health crisis, the acute illness management had been obvious primary focus. However, the growing evidence of post-COVID conditions warrants the shift in focus and consider COVID-19 as an alternative diagnosis if that is co-related.

## Conclusion

The prevalence of post-COVID-19 symptoms among the study population is found to be high. The main symptoms from our study and other similar studies include anxiety, fatigue, loss of appetite, shortness of breath, muscle pain and headache. Patients who smoked, had hypoxia during acute illness, had presence of chronic lung disease or neurological disorder had higher odds of getting post-COVID-19 conditions. Citing the findings, health system should focus upon the management and recording of such conditions for further evidence.

## Data Availability

The data underlying the results presented in the study are available from Curative Service Division, Department of Health Services.

## Acknowledgment

We would like to express our sincere gratitude to Dr. Roshan Pokharel, the Secretary of the Ministry of Health and Population, and Dr. Rajesh Sambhajirao Pandav, Country Representative for WHO, Nepal for their overall leadership and support in developing this research manuscript. We are also deeply grateful to all individuals who are directly or indirectly involved in the entire process from conceptualization to the publication stage including those whose details are included in the study.

